# A qualitative exploration of the psychosocial factors affecting antiretroviral therapy adherence among HIV infected young adults in Eastern Uganda

**DOI:** 10.1101/2023.02.04.23285423

**Authors:** Easton Twekambe, Joshua Epuitai, Enid Kagoya Kawala, Vian Namanya, Emmanuel Tiyo Ayikobua, Samuel Baker Obakiro, Agnes Napyo, Kenedy Kiyimba

## Abstract

**Background:** The use of anti-retroviral therapy (ART) in management and prevention of HIV/AIDS epidemic is a globally accepted strategy. In Uganda, despite the efforts to increase uptake of ART, adherence remains a huge challenge. This study, therefore, was conducted to explore psychosocial factors which influenced non-adherence to ART among young adults in Eastern Uganda.

**Methods:** This was an exploratory qualitative study conducted at the ART clinic of Mbale Regional Referral Hospital. A total of 35 in-depth interviews with young adults who had defaulted from taking ART drugs were conducted. Also, five key-informant interviews were conducted among the healthcare workers in the hospital. Thematic analysis approach was followed to analyze the data using NVIVO software (version 11).

**Results:** Non-adherence to ART was perceived to result from poor social support, poor coping mechanisms, unpredictable and busy work schedules, poverty, incompatible religious beliefs and practices. The poor social support factors included poor caregiver support, non-disclosure of HIV status, stigma and discrimination while factors linked to poor coping mechanisms included alcohol and substance abuse, psychosocial stress, depression, forgetfulness and feelings of self-hatred. Poverty limits access to treatment and basic needs including transportation to health facilities. Long waiting time at the ART clinic was the healthcare system factor which was thought to cause non-adherence among young adults.

**Conclusion:** Non-adherence to ART at Mbale Regional Referral Hospital in Eastern Uganda is attributed to various psychosocial factors such as stigma and discrimination, mental health problems, work-related problems and low socio-economic status, religious beliefs and poor knowledge, long waiting time in ART clinic and poor family support. Psychosocial counselling should therefore target the perceived causes of ART non-adherence so as to improve the adherence /compliance to ART.

## 1.0 Background

HIV/AIDS is still an epidemic with devastating consequences to humanity worldwide. Globally, an estimated 38.4 million people were living with Human Immune Virus (HIV) in 2021 of which 54% (20.6 million) of the population were in Southern and Eastern Africa [1]. Uganda is one the countries in sub-Saharan Africa with the high burden of HIV infection [2]. In 2021, the prevalence rate of HIV among adults aged 18-49 in Uganda was 5.2% which translated to about 1.3 million people [2]. Uganda is a predominately young population with significant public health implications given that the risk of new infections is highest among the young adults [3]. In Uganda, 37% of all the new HIV infections were among the young adults aged 15-24 years in 2020 [4].

The United Nations AIDS (UNAIDS) has set an ambitious 95-95-95 target to eliminate HIV/AIDS epidemic by 2030 [5]. The 95-95-95 strategy is where 95% of the population are aware of their HIV status, 95% of HIV positive clients are on antiretroviral treatment (ART), and subsequently 95% of HIV positive clients on ART are able to achieve HIV viral load suppression [5]. In Uganda, in 2020, 89% of people living with HIV were aware of their HIV status, 82% of people living with HIV were on ART, and 78% of people living with HIV had viral suppressed loads [2]. Consequently, Uganda is far from meeting the 95-95-95 target [2].

Optimal elimination of HIV transmission and viral load suppression requires strict adherence to ART [6]. The risk of treatment failure is likely to occur following failure to adhere to the lifelong ART [7-9]. Young adults are more likely than adults to miss taking ART drugs [6]. A previous study among youths showed that 37% of the youths living with HIV refused or missed taking ARVs for a period of one month in Rwanda [10], while another study in Uganda reported that 90% of the youths had more than 95% adherence in taking ART drugs [11]. Poor adherence to ART was attributed to poverty, stigma, non-disclosure, discrimination, pill burden, fatigue, depression and side effects [11-17]. Short waiting time, counselling, supportive healthcare workers, peer support and provision of food and transport was found to facilitate good adherence practices among the youth [11]. Previous studies explored adherence in the context of married couples [18], while a few studies among the young adults were quantitative in nature [11]. In this study, a qualitative research design was used so as to further explore and understand the psychosocial factors which hindered adherence to ART among young adults living with HIV.

## 2.0 Methods and Materials

### 2.1 Study design and setting

This study used a phenomenological qualitative study design to explore psychosocial factors which influenced non-adherence to ART among young adults at the ART clinic of Mbale Regional Referral Hospital (MRRH). MRRH is the biggest hospital in Eastern Uganda with a 400-bed capacity serving over 16 districts in Eastern Uganda. The ART clinic at the hospital offers a wide range of services which include health education, HIV counseling and testing, ART dispensing, laboratory services, clinical assessment, ART adherence counseling and psychosocial support.

### 2.2 Study Participants

HIV positive young adults from age 19-35 years who were receiving ART in the hospital and were non-adherent to ART were recruited in the study. Participants who defaulted on ART with adherence of less than 90% in the past three months were selected and were considered to be non-adherent to ART.

The study also included ART clinic staff purposively selected since they worked so closely with the young adults. These clinic personnel included a clinical officer, a nurse who was the in-charge of the ART clinic, a social worker and two primary health care counselors. All the participant Identifiers were not disclosed nor known to anyone outside the research group.

### 2.3 Sample size calculation

The sample size for this study was based on the principle of data saturation when no new information was collected from the interview[19]. In this study, data saturation was reached when a total of 35 in-depth interviews and five key informant interviews were conducted.

### 2.4 Data collection

In-depth interviews (IDIs) were used to collect data from study participants. The interviews were conducted in Luganda, Lugishu and English. The IDIs were conducted by ET, a nursing student in the final year of study and a native speaker of Luganda. The interviews were conducted in a quiet room with adequate privacy. The IDIs with the participants lasted a duration of about 30-45 minutes. A topic guide with open-ended questions were used during the IDIs. The questions asked included what challenges hindered these clients from adhering well to the recommended lifestyle of people living with HIV while at school, home, work places, health facility, any illicit drugs or alcohol use by these clients, feelings of self-hatred, homicidal and suicidal ideas. In addition, social demographic information including age, sex, marital status, level of education, religion, and time the client has spent on ART were collected. Key informant interviews were conducted to understand the perceived psychosocial factors for ART non-adherence among young adults from the lens of experts involved in ART. Key informant interviews, lasted a duration of about 30-45 minutes, were conducted in English using an interviewer guide. The interviewer guided contained questions such as what challenges hinder these clients from adhering well to the recommended lifestyle for people living with HIV while at school, home, work places, health facility, any illicit drugs or alcohol use by these clients, report feelings of self-hatred, homicidal, suicidal ideas or any other mental illnesses. Method triangulation, through use of IDIs and key informant interviews, ensured rigor of the data collected in the study. Audio recording of the interviews was done.

### 2.5 Data analysis

The audio recording were transcribed word to word from Luganda and Lugishu to English by a trained native Luganda speaker (ET and EK). NVIVO version 11 plus was used in the data analysis and processing [20]. Brauna and Clarke thematic analyses was used to analyze the data [21]. The transcript was read for several times to get familiar with the data [21]. A multidisciplinary team of researchers including a social worker, nurse, counsellor and a pharmacologists) were involved in data analysis. The codes generated were discussed by the team to ensure that the codes were grounded and consistent with the data. The interviews were then read by the social scientist (EK) who randomly selected and independently coded three interviews [21]. Codes, sub-themes, and themes were used to describe the data [21].

### 2.6. Ethical clearance

Ethical approval from Mbale Regional Referral Hospital Ethics and Research Committee (reference number: MRRH-2021-82) was obtained. Informed consent was obtained from individual participants, while confidentiality and privacy were strictly followed; the participant Identifiers(IDs) were not known to anyone outside the research group. The study was conducted during COVID-19 pandemic, and as such, compliance to standard operating procedures were strictly maintained during data collection.

## 3.0 Results

### 3.1 Study participant demographics

A total of thirty-five young adults living with HIV were enrolled in the study. The participants included those who had an adherence of less than 90% in the period of the last three months (table 1). The work experience of key-informants ranged from two to eight years.

**Table 1:** Description of Socio demographic characteristics of participants from IDIs.

| Demographics | Frequency(N=35) | Percentage (%) |
| --- | --- | --- |
| **Age** |  |  |
| 19-24 | 14 | 40.0 |
| 25-30 | 8 | 22.9 |
| 30-35 | 13 | 37.2 |
| **Gender** |  |  |
| Female | 20 | 57.1 |
| Male | 15 | 42.9 |
| **Level of education** |  |  |
| Primary or no education | 14 | 40.0 |
| Secondary | 16 | 45.7 |
| Tertially | 5 | 14.3 |

### 3.2: Psychosocial factors perceived to influence ART non-adherence

Five themes embodied the perceived psychosocial of ART non-adherence: poor social support, religious beliefs, poor coping mechanisms, poverty and unpredictable schedules, and health system related factors (table 2).

**Table 2:**
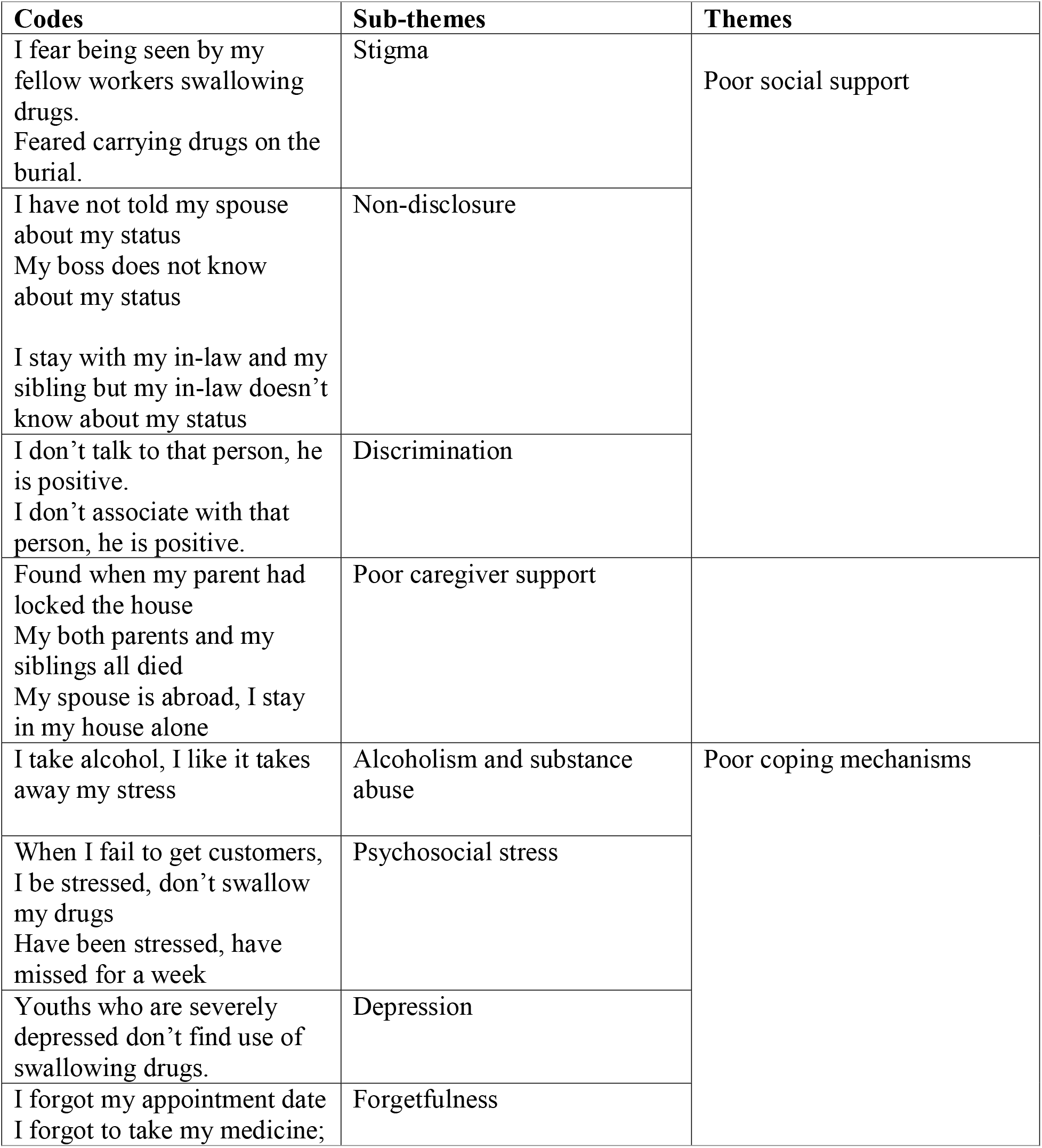
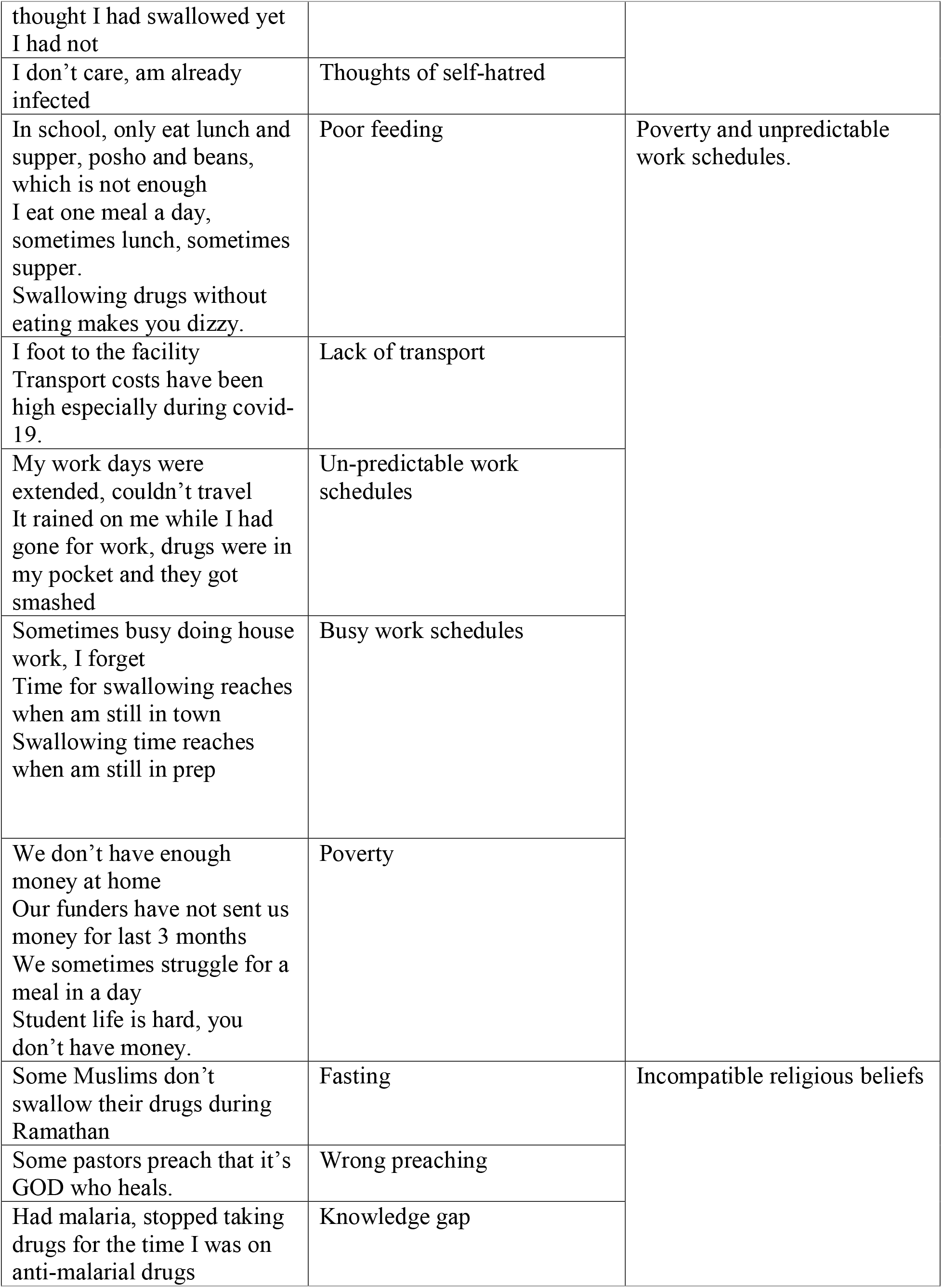
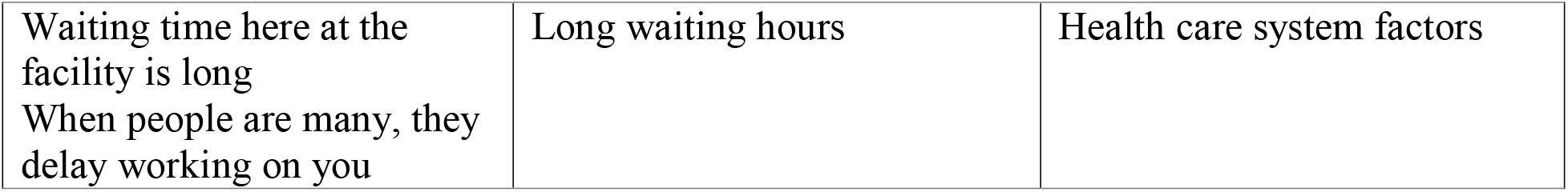
psychosocial factors influencing ART adherence.

#### 3.2.1. Theme1: Poor social support

##### 3.2.1.1 Stigma

Some clients attributed missing swallowing their ART medicines to fear of being seen by friends, partner, relatives, and workmates. Fear of being seen taking medications was mostly expressed by students in school, and participants who resided with their fellow colleagues.

*“I am a housekeeper in a hotel, we sleep in a dormitory together with my other colleagues, I hide and swallow my medicine like at 9:00 am such that my workers don’t see me”, (IDI-15, with inconsistence in time of swallowing)*

*“I am a track driver, sometimes I get a customer in my lorry and time for swallowing reaches, I only have to stop the lorry, move out as if I am going to urinate as I swallow my tablet without the customer seeing me” (IDI-09, missed about 5 times in last 3 months)*

The young adults in schools feared swallowing or picking drugs from their suite cases when fellow students were seeing them.

*“I am a boarding student, I wake up very early in the morning, put my medicine in my pocket, swallow them at around 10 am when I feel I am in a private place”. (IDI-25, missed 5 times while at school)*

*“I missed about 5 times while I was at school because I feared fellow students to see me picking my drugs from my suitcase”, (IDI-17)*

*“At our school I swallow my drugs from the sickbay but it’s near a boy’s dormitory. Fellow students keep wondering why I have to go to the sick bay on a daily basis”, (IDI-32 missed 10 tablets in 3 months)*

Some clients reported that events that occur abruptly make them fail to adhere well to taking their drugs and honor appointments. Abrupt burials were perceived to cause some clients to miss taking their ART treatment because of stigma and fear from being seen taking drugs by others

*“I had gone for the burial, I feared carrying my medicines”, (IDI-09, missed 3 tablets)*.

##### 3.2.1.2 Non-disclosure

Non-adherence to ART medications were attributed to failure of the clients to disclose their HIV status to their partners, relatives and neighbors, and the fact that they were taking their ART medications. Because of non-disclosure, the clients could not take their medications in the presence of the relative, and consequently were not consistent in taking the medications at the required time.

*“I have so far spent 6 months on treatment, I have not told my spouse about my status, I hide and swallow my medicine when he is not seeing me or wait and swallow when he is not around” (IDI-16, reported inconsistences in time of swallowing)*

*“I stay with my sibling together with my in-law at their home. My in-law doesn’t know about my status and we don’t want him to know about it. I swallow my medicine at 9:00 am and if he delays to go for work, I wait until he goes for work then swallow my medicine such that he doesn’t see me*.*” (IDI-17)*

##### 3.2.1.3 Discrimination

Some key informants perceived that HIV positive clients were discriminated against by the society, which the key informants thought promoted poor adherence practices among the clients. Clients who were discriminated were perceived to fail to take their ART medications because the lack of social approval led to poor coping mechanisms including self-hatred.

*“I don’t talk to that person, he is positive”, “I don’t associate with that person, he has HIV”, (KI-03, counselor for 3 years at the clinic)*.

#### 3.2.2 Theme 2: Poor coping mechanisms

##### 3.2.2.1 Stress

Work-related stress was perceived to cause non-adherence to ART medications, while stress related to financial insecurities was attributed to cause poor adherence practices in some of the clients.

*“I rent in town and my funders have spent 3 months without sending us rent for the last three months. This has always stressed me and so far, I have skipped swallowing for the whole of last week”, (IDI-14)*

*“I work in a saloon here in town, sometimes I fail to get customers for a day, I go back home when I am stressed and fail to swallow my medication”, (IDI-07)*

##### 3.2.2.2. Forgetfulness

Poor timing of taking ART drugs, missing taking the drugs, and missed appointments were partly attributed to forgetfulness by the clients.

“*I missed one tablet in the last 3 months, I forgot to take, me thought I had taken yet I had not”, (IDI-10)*

*“I was supposed to return yesterday for more drugs but I forgot the return date”, (IDI-04, missed appointment)*.

##### 3.2.2.3. Depression

Key informants reported that clients who were newly diagnosed with HIV infection lost hope and sometimes were severely depressed which made the clients to stop taking ART medications.

*“Severely depressed clients don’t find the use of swallowing drugs until they are given various sessions of psychosocial counselling”, (KI-03, counselor for 3 years at the clinic)*.

##### 3.2.2.4 Alcoholism and substance abuse

Non-adherence to ART was attributed to alcohol intake as it affected the cognition of the clients and made the clients to forget to take their medicines or even delay to return back home to take their drugs.

*“I take alcohol because I like it and it takes away my stress. However, it has made me miss several times since I forget. Sometimes I deliberately refuse to take these medicines whenever I am drunk. I missed very many times because I always get drunk”, (IDI-14)*

##### 3.2.2.5. Thoughts of self-hatred

Poor adherence was attributed to ill-thoughts about HIV infection that made some clients miss swallowing the medication since they did not care, they were already infected

*“I don’t care, I am already infected”. (IDI-14, missed very many times)*

#### 3.2.3 Theme 3: Poverty and unpredictable work schedules

##### 3.2.3.1 Poverty

Nutrition and good feeding is important for HIV clients who are on ART medications. Lack of money to afford basic needs such as food was perceived to make clients to skip or miss taking the ART medications. Clients were not able to honor their appointments to come to the facility to pick their ART because of the lack of transport fares. Clients claimed that they failed to raise money for transport to keep appointment dates during COVID-19 especially with increased taxi fares during covid-19 lockdown where transport fares were doubled.

*“I stay with my parent, we don’t have enough money, sometimes we sleep without having supper”, (IDI-21, missed once in last 3 months)*

*“Our funders have spent 3 months without sending us money, I am stressed that’s why I have been skipping”, (IDI-14, missed very many time)*

*“We don’t have enough funds at home, we even struggle to have one meal a day”, (IDI-08)*

*“Covid-19 restrictions have made transport fares high, if I delay at the facility, I end up using 15000UGX to 20000UGX (4*.*24$ to 5*.*65$) on a boda-boda which is very expensive for me”, (IDI-03, missed appointment date)*

*“Student life when you have nothing to eat is very hard, it’s harder to swallow drugs when you are dizzy and hungry”, (IDI-13*.)

##### 3.2.3.3 Poor feeding

Related to poverty, failure to adhere to ART medications at home was attributed to lack of food. The clients appreciated that ART drugs required good feeding at least breakfast, lunch, dinner and supper and also the need for adequate fluid intake. s. Some clients claimed that they felt well when the drugs were taken using good juice and not with plain water. Respondents from secondary schools especially those in boarding reported poor feeding practices including lack of a balanced diet at schools and inadequate breakfast while some ate only lunch and supper with poor quality of food. Clients reported that was difficult for them to take the ART drugs when they were hungry since the drugs would make one dizzy.

*“It’s hard for me to find what to eat while I am in this town, I eat breakfast and one more meal for a day, and sometimes I do only lunch and sometimes do only supper*.*” (IDI-14, missed very many times)*

*“I could miss the drugs especially when I have nothing to eat”, (IDI-07)*

*“I am always weak at school, I only feed on posho and beans which is always little for me”, (IDI-25)*

*“I am in boarding school, we always have only 2 meals a day lunch and supper where we only eat posho and beans, I am not always satisfied that’s why I end up swallowing poorly”, (IDI-11)*

*“Sometimes I have no food these days and these drugs make me feel dizzy when I take them on an empty stomach”, (IDI-12)*

*“I stay with my parent who has been on treatment for so long, my parent died, sometimes I sleep without having supper, I only ensure that my young baby has eaten”, (IDI-21)*.

##### 3.2.3.2 Unpredictable work schedules

Unpredictable work schedules and prolonged duration of work away from home was reported to cause some clients to miss taking their medications. These clients were not very sure of the number of days they would spend at their workplaces thus ended up missing since their work places were very far thus could not pick tablets for extra days.

*“I missed about 6 tablets in the last 3 months because whenever I get work, my work days are extended yet home is always far for me to go back and I pick my medicines”, (IDI-34)*

*“I had gone for work, carried about 5 tablets in my pocket that I would use during that period, it rained on me and they all smashed, couldn’t come back home to pick more drugs because it was very far,” (IDI-09, missed 5 tablets)*

##### 3.2.3.4. Busy schedules

Some clients attributed the poor timing and failure to swallow the drugs well to busy schedules at home. They ended up getting caught up in other activities thus delaying to swallow the drugs well and thus poor adherence. Busy schedules also affected young adults who were in school where a lot of time was spent in class and preps and poor time keeping of school activities meant that the time of taking the drugs collided with school activities.

*“I am sometimes busy doing house work at home, and thus my time has not been constant 10:00pm”, (IDI-22)*

*“My time for taking, 9:00 am, reaches when I am already needed in town, thus have to take my medications before that time such that I don’t miss”, (IDI-18)*

*“I am in a boarding school, sometimes I miss coming for review because of tight school programs and I have to send someone to come and pick for me the medicine”, (IDI-03)*

*“In school, my time 10:00pm collides with prep time, sometimes I am still in class or still discussing with my classmates”, (IDI-22, reports inconsistences in timing)*

#### 3.2.4 Theme 4: Incompatible religious beliefs

Key informants attributed religious belief to cause clients to miss taking their ART medications. The staff reported that some religious leaders were a burden to good adherence of their clients and also their religious affiliations.

*“Some pastors whom they believe in lie them that God will heal them even when they don’t take drugs, and thus clients will end up stopping to swallow their medications well waiting for miracles”. (KI -05)*

*“Some Muslims on ART stop swallowing their drugs during Ramathan. They miss due to fasting”, (KI-04)*

##### 3.2.5.3 Knowledge gap

Some clients missed swallowing drugs due to lack of adequate knowledge regarding the need to continue taking medication regardless of their health status. Some participants stopped taking ART medications when they were sick and on other treatments, and resumed taking ART medications after completing taking medications of other illness.

*“I was on treatment for malaria, I decided to finish drugs for malaria and resume ARVs later”, (IDI-26, missed 6 tablets in last 3 month)*.

#### 3.2.5. Theme 5: Family support

##### 3.2.5.1. Poor family support

Some respondents who lacked caring marriage partners, parents and relatives were finding problems in adhering well to ART. Clients sometimes forgot to swallow their medicines and were not reminded by their partners leading to inconsistent timing and or missing to take the drugs.

*“Missed once, returned home very early in the morning, found when my parent had locked the house, couldn’t find my drugs since they were locked inside our house”, (IDI-21)*

*“My parents both died together with my siblings and I am the only one living, I stay with my relative and his spouse, sometimes I fail to get what to eat”, (IDI-17*.)

*“My spouse went and he works abroad, I am the first borne but I stay alone in my house, I left my parents. Sometimes I forget swallowing”, (IDI-09)*

Poor adherence among the married young adults was attributed to unfaithful partners. Some women claimed that whenever their spouses failed to fulfill their family obligations of providing for the family, they ended up stressed and missed taking the pills for some days.

*“I stay together with my spouse and my two children who are all on treatment. My spouse always cheats on me, we end up lacking money, I am always stressed about it”, (IDI-24)*

## 4.0 DISCUSSION

This study was conducted to explore the psychosocial factors that hindered adherence ART among HIV positive young adults. Non-adherence to ART was perceived to result from stigma and discrimination, mental health problems, work-related problems and low socio-economic status, religious beliefs and poor knowledge, healthcare system factors and poor family support. Addressing these psychosocial factors hindering adherence to ART would promote increased compliance to treatment and hence reduced morbidity and mortality form HIV/AIDS. Stigma and discrimination were described to affect all young adults on care and it was among the most common reasons of failure to swallow the pills while at school, work places, and homes. Stigma is due to perceived unbearable burden for taking medication for life which lessens the self-confidence of the clients leading to non-adherence [11, 15]. Stigma in the healthcare facilities manifests in form of internalized fear of being seen in ART clinic, impatience to wait, and rush in picking the drugs [15]. A study done in Northern Uganda among young adults also reported stigma as the main cause of non-adherence to ART [11]. Similar findings have been reported in Myanmar, where enacted stigma and internalized stigma were associated with worse ART adherence [13]. Stigma and discrimination restrict the freedom of HIV positive in taking their ART pills in the presence of other people resulting in delaying, poor timing and non-adherence to ART. Stigma remains a stumbling block to good adherence and its impact is not only reflected on the recipients of care but also on their care takers, communities, and the entire health care system [15]. The consequent poor counselling and inadequate holistic health assessment of the client worsens adherence practices but may also contribute to ART failure [15].

Disclosure of HIV status has remarkable benefits in promoting drug adherence among other social benefits[22, 23]. In our study, clients who had not disclosed their HIV status found it difficult to ask permission from their superiors to allow them to go and pick their medication from the health facility. Missed appointments, poor timing and skipping to take their medication was noted among students who had not disclosed their HIV status as they were unable to seek permission, special consideration in terms of meals, and receiving adequate social support from the school manned clinic. Similar findings were cited in Ghana where non-disclosure led to low adherence on ART [12]. Non-adherence was significantly more common among clients who had not disclosed their HIV status to their partners, a finding which was consistent with study findings [12]. Therefore, clients who have not disclosed their HIV status should be counselled on the role of disclosure in improve adherence to ART.

Consistent with previous studies [14, 16, 24], failure to adhere to ART was attributed to mental health problems including stress, depression, feelings of self-harm, poor coping mechanisms and use of illicit drugs. In Malawi, young adults who had depressive symptoms were significantly more likely to default on their ART medication [25]. Like in previous studies [14, 24], newly diagnosed HIV positive clients were more likely to resort to the use of alcohol and other illicit drugs to cope with the associated psychosocial stress. Alcohol and substance intake affects social cognition and lowers social inhibition of the clients leading to forgetfulness, irrational behaviors and decisions which consequently comprise the ability of HIV positive clients to adhere to their medications [14]. Promoting adherence requires strong psychosocial counselling services to screen, identify, and appropriately manage clients who are likely to start using drugs and as well clients who are already abusing drugs.

Previous studies have indicated that long waiting time in ART clinics was significantly associated with poor ART drug adherence [26, 27]. A study revealed non-adherence to ART was attributed to long waiting time in 24% of clients [27]. Consistent with previous studies [28], long waiting times and high volume of clients in ART clinics meant that some clients did not honor their appointments, while some clients spent so much time in the clinic, and chose to leave the clinic without receiving their drugs. The high volume of patients relative to the few healthcare workers compromises on the quality of client-healthcare worker interaction, privacy, poor quality of care, and failure of the healthcare workers to identify issues of ART adherence among clients [28]. Measures should be developed to reduce long waiting times in ART clinics so as to reduce issues of non-adherence to medications and consequent treatment failure

Consistent with findings from previous studies[28, 29], poverty, and low-socio economic status contributed to the ART non adherence among HIV young adults. In a scoping review of the factors contributing to ART adherence, logistical and financial support to HIV positive clients were found to facilitate adherence to ART medication [30], which also supports our findings. Non-adherence from lack of money was attributed to failure of the clients to afford basic needs such as food and transport fares to cater for additional demands of taking ART medications. COVID-19 restrictions were attributed to cause poor ART drug adherence related to financial insecurity from closure of business, non-paid leave, laying off workers, and doubling of transport fares. Financial hardships together with poor quality of meals at schools meant that while in school students were not able to buy additional food to supplement their feeding and consequently cope with the demands of taking ART medications. This underscores a need for schools to provide special support to HIV positive clients in school through provision of special meals in order to promote drug adherence.

Some religious beliefs were perceived to be associated with poor adherence to ART medication. Some Muslim clients were implored to stop taking their medication during Ramadan, while some pastors gave false hope to their followers that God would heal them even when they do not take medication. The findings of our study were in agreement with a study findings [31, 32] which cited poor ART adherence was related beliefs in faith healing, alternative traditional medication, and perceptions that HIV was caused by witchcraft. However, this was contrary to the findings in Ghana, where religious beliefs empowered the clients to ignore stigma, become resilient and motivated to take their medications because of their beliefs that God approved the use of ART thus promoting good adherence [33].

## 6.0 Conclusion

Poor timing of taking medication, missed appointments to pick medications, and defaulting in taking ART medication was perceived to result from stigma and discrimination, mental health problems, work-related problems and low socio-economic status, religious beliefs and poor knowledge, long waiting time in ART clinic and poor family support. Mental health problems including stress, alcohol and substance use, and depression were perceived to negatively affect drug adherence. There is need for continuous counselling of young adults living by the Healthcare providers on, mental health, ART adherence, healthy lifestyle, and prompt management of ART side-effects.

## Data Availability

The datasets used and/or analyzed during the current study is available at https://doi.org/10.5061/dryad.ns1rn8pxp.

## Abbreviations

AIDS: Acquired Immunodeficiency Syndrome
ARV: Antiretroviral drugs
ART: Antiretroviral therapy
HIV: Human immunodeficiency Virus
MRRH: Mbale Regional Referral Hospital
HMIS: Health Management Information System
IDI: in-depth interviewees
KI: key informants

## Declarations

**The authors have no conflict interest to declare**

## Ethics approval and consent to participate

Ethical approval was sought from the Research and Ethics committee of Mbale Regional Referral Hospital. Each participant was made aware that participation is voluntary and that they can opt out of the study at any stage without any penalty. The information shared with the investigator was kept confidential and private by restricting any data access. To maintain privacy, names and any other individual identification were not recorded but unique in-depth interviewee identification numbers and key informant numbers were used in participant identification. Informed consent was sought before data collection from a participant and participants were asked to append their signatures on the consent form. All participants were well educated about the objectives of the study, any risks and benefits.

## Consent for publication

Not applicable

## Availability of data and materials

The datasets used and/or analyzed during the current study are available from the corresponding author on reasonable request.

## Authors’ contributions

ET conceived the idea, JE, EKK contributed in data collection, data analysis and interpretation, VN, EAT, SBO, AN, KK participated in writing of the manuscript. All authors read and approved the final manuscript.

## Acknowledgements

We are grateful to Mbale Regional Referral Hospital, the key informants and the study participants for their valuable input in the study

